# GLAPAL-H: Global, Local, And Parts Aware Learner for Hydrocephalus Infection Diagnosis in Low-Field MRI

**DOI:** 10.1101/2025.05.14.25327461

**Authors:** Srijit Mukherjee, Kelsey Templeton, Starlin Tindimwebwa, Ivy Lin, Jason Sutin, Mingzhao Yu, Mallory Peterson, Chip Truwit, Steven J. Schiff, Vishal Monga

## Abstract

**Objective:** The study aims to develop a method for differentiating between healthy, post-infectious hydrocephalus (PIH), and non-post-infectious hydrocephalus (NPIH) in infants using low-field MRI, which is a safer, low-cost alternative to CT scans. The study develops a custom approach that captures hydrocephalic etiology while simultaneously addressing quality issues encountered in low-field MRI.

**Methods:** Specifically, we propose GLAPAL-H, a Global, Local, And Parts Aware Learner, which develops a multi-task architecture with global, local, and parts segmentation branches. The architecture segments images into brain tissue and CSF while using a shallow CNN for local feature extraction and develops a parallel deep CNN branch for global feature extraction. Three regularized training loss functions are developed — one for each of global, local, and parts components. The global regularizer captures holistic features, the local focuses on fine details, and the parts regularizer learns soft segmentation masks that enable local features to capture hydrocephalic etiology.

**Results:** The study’s results show that GLAPAL-H outperforms state-of-the-art alternatives, including CT-based approaches, for both Two-Class (PIH vs. NPIH) and Three-Class (PIH vs. NPIH vs. Healthy) classification tasks in accuracy, interpretability, and generalizability.

**Conclusion/Significance:** GLAPAL-H highlights the potential of low-field MRI as a safer, low-cost alternative to CT imaging for pediatric hydrocephalus infection diagnosis and management. Practically, GLAPAL-H demonstrates robustness against quantity and quality of training imagery, enhancing its deployability. The code for this work is available here: https://github.com/mukherjeesrijit/glapalh.

## I. Introduction

Hydrocephalus, a condition characterized by abnormal cerebrospinal fluid (CSF) accumulation within the brain’s ventricular system, poses a significant challenge in pediatric neurosurgical care globally. In sub-Saharan Africa, postinfectious hydrocephalus (PIH) accounts for over 50% of pediatric hydrocephalus cases, while non-postinfectious hydrocephalus (NPIH) stems from diverse etiologies such as hemorrhage and congenital malformations [1–4]. These distinct etiologies manifest unique anatomical pathologies on neuroimaging, with infectious cases often presenting with calcifications (in CT scans), abscesses, loculations, and intraventricular debris. Both PIH and NPIH typically result in ventricular dilation, brain tissue edema, and increased head circumference. [4, 5] The structural alterations in hydrocephalic (PIH, NPIH) brains deviate significantly from healthy brains, as depicted in Fig. 1 (A). Hydrocephalus is characterized by markedly enlarged and distorted ventricles, leading to compression of surrounding brain tissue and potential neurological deficits and developmental delays if left untreated. Accurate differentiation between healthy, PIH, and NPIH infants is critical for optimal patient management, as surgical intervention may need to be delayed in cases of active brain infection until appropriate antimicrobial treatment is administered [6–8]. This distinction underscores the importance of developing robust diagnostic tools and treatment strategies tailored to the specific etiology of hydrocephalus. For infants with hydrocephalus, diagnostic imaging typically involves computed tomography (CT) scans due to their accessibility [9]. But, CT scans expose patients, especially children, to ionizing radiation, increasing the risk of brain tumor induction [10, 11]. Recent studies [12–15] have highlighted the effectiveness of low-resolution brain imaging in managing infant hydrocephalus. This aligns with the increasing adoption of portable low-field MRI systems, like the Hyperfine MRI scanner, which provides imaging solutions in resource-constrained environments. However, developing an effective classifier for low-field MRI images presents significant challenges due to their low resolution and quality issues. Traditional computer-aided diagnosis systems (CADs) typically require learning representative features by dictionary learning [16] or manual extraction of features from images to feed subsequent classifiers such as support vector machines or decision trees [17, 18]. These classical methods often struggle with high-dimensional data and require substantial domain expertise for feature engineering. The increasing complexity of medical imaging data has led to a shift towards deep learning approaches, particularly Convolutional Neural Networks (CNNs), which can automatically extract hierarchical features from raw image data. Deep learning models offer improved accuracy and efficiency across diverse medical applications. Both 2D and 3D CNNs have been effectively applied to various tasks, including brain tumor detection, lung nodule/cancer classification, COVID-19 detection, skin cancer classification, and other medical diagnoses [19–37]. Despite the inherently black-box nature of many deep learning models, recent efforts, such as a survey by Guan et al. [38], have emphasized domain-adapted learning, highlighting the significance of non-black-box AI approaches. Notably, the Brain Attention Regularizer Network (BAR-Net) [39] has demonstrated high accuracy in detecting infections in hydrocephalus using CT images by leveraging a domain-enriched attention-learning framework.

**Fig. 1.**
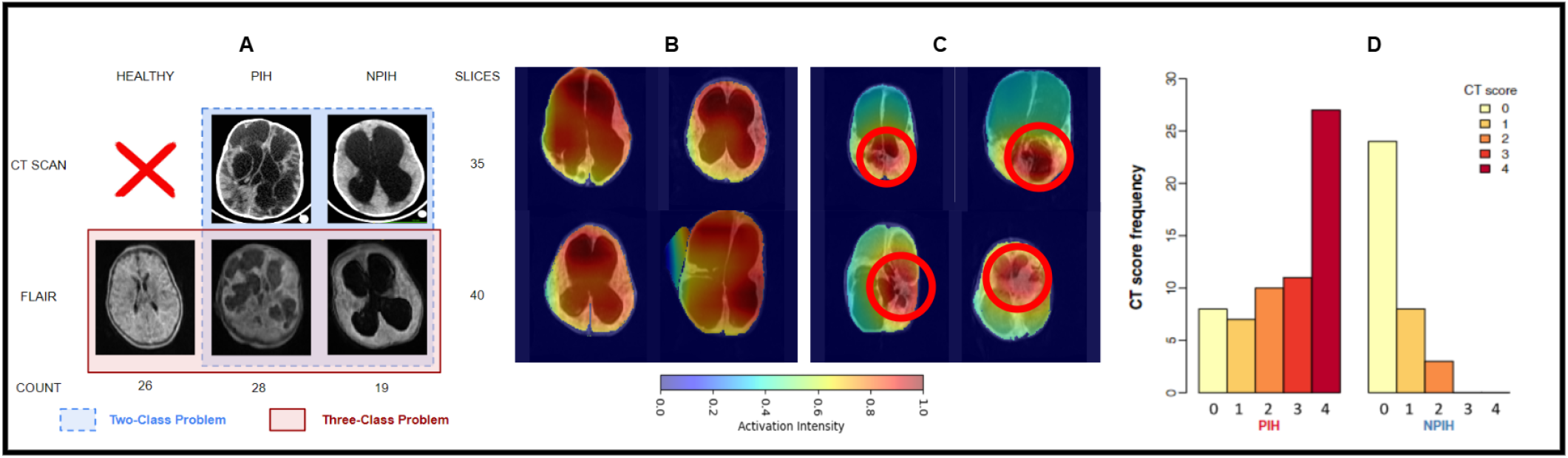
(A) Comparison of CT and low-field MRI for PIH, NPIH, and healthy infants, with slice counts and label distribution. Note the quality difference between high-quality CT and low-field FLAIR (Fluid Attenuated Inversion Recovery) MRI. CT scans are omitted for healthy infants due to radiation risk. (B, C) BAR-Net [39] activation maps for low-field MRI: (B) global activation across the brain, (C) localized activation (red circles) in brain tissue or CSF, inspiring segmentation-based feature extraction and classification. (D) From [5], reprinted with permission from AAAS, the figure shows preoperative CT scan scoring by two experienced neurosurgeons, assigning points for loculations, debris, calcification, and abscess formation.

Machine learning-based medical image classification for infection diagnosis using low-field MRI remains a relatively unexplored area, with notable exceptions including artifact detection [40] and malaria detection using traditional non-deep learning algorithms [41]. Recent research has focused on leveraging deep learning to address various challenges associated with low-field MRI like accelerating the imaging process [42], improving image reconstruction [43, 44], enhancing overall image quality [45], applying super-resolution techniques [46], and developing tailored segmentation algorithms [47]. These advancements demonstrate the potential of combining deep learning with low-field MRI to overcome its limitations. Recent work [48–63] have demonstrated the benefits of multi-task learning for medical image classification, where the auxiliary task involves segmentation of an *apriori* known and well-defined artifact such as tumor, lesion, etc.

The above methods mostly rely on high-quality imaging modalities such as CT, MRI, Ultrasound, and X-ray. Unlike well-defined, localized disease-specific features in the aforementioned methods, post-infectious hydrocephalus presents unique challenges due to diverse, distributed infection regions in infant brains as evident from Fig. 1 (A), further complicated by low-resolution low-field MRI. Hydrocephalic infant brain imagery lacks *apriori* information on infection locations in brain tissue or CSF. This distributed nature of infection markers, combined with resolution limitations, makes accurate diagnosis particularly challenging. Nevertheless, the success of segmentation-based classification in other medical imaging tasks suggests promise for addressing this problem.

We develop a novel segmentation-guided classification approach for hydrocephalic infection diagnosis. Unlike [51, 56, 60], which exploit *apriori* knowledge of the shape and expected location of tumors, we perform a parts-based segmentation on the brain image into brain tissue and CSF guided by custom-designed regularizers. This is motivated by the fact that hydrocephalic infection is dictated by etiologies present in brain tissue, CSF, and their interplay. Our proposal is hence termed GLAPAL-H: Global, Local, And Parts Aware Learner for Hydrocephalus Infection Diagnosis in low-field MRI. Our key contributions are as follows:

1. **Multitask architecture with global, local, and segmentation branches:** The class activation maps for low-field MRI Fig. 1 (B, C) from past work [39] in hydrocephalic infection diagnosis, demonstrate that infection may be contained in brain tissue/CSF parts or distributed globally. This motivates GLAPAL-H’s incorporation of three fundamental branches: global, local, and parts (brain tissue, CSF) segmentation. The global branch extracts features from larger receptive fields, while the shallow local branch focuses on smaller regions to spatially localized infection signs. The segmentation branch divides the brain region into two parts: brain tissue and CSF, guiding the local branch. This multitask approach is designed to capture both widespread and localized features of hydrocephalus in low-resolution MRI images, addressing the unique challenges presented by the diverse and distributed nature of infection regions in infant brains.
2. **Problem inspired Global, Local, and Parts Regularizers:** We leverage insights from Fig. 1 (B, C) regarding global features (large-scale global assymetry) and from Fig. 1 (D) regarding local features (septation, abscesses, debris) in hydrocephalus diagnosis. To ensure complementary learning between branches, GLAPAL-H employs three unique regularizers. The global regularizer extracts high-level global features, while the local regularizer along with the shallow local branch facilitates the extraction of fine-detailed features in local contexts. The parts regularizer utilizes segmentation features and ground truth masks to create *learnable soft masks* that in turn help the GLAPAL-H network to extract the relevant discriminative features from brain tissue and CSF separately.
3. **Hydrocephalus low-field MRI Adaptation:** Despite the inherently lower resolution and tissue contrast of low-field MRI [13], [64], they have been shown to contain reliable information for disease diagnosis, specifically hydrocephalus [12]. Central to this diagnostic capability is that discriminative cues for hydrocephalic infection are present in brain tissue, CSF, or both. The multi-branch design of GLAPAL-H and its parts-based approach is designed to adapt to quality imperfections of low-field MRI, so relevant diagnostic information may be extracted from any of the branches.
4. **Experimental Validation and Insights:** Our work introduces and exploits a *new dataset* of low-field Fluid Attenuated Inversion Recovery (FLAIR) MRI ^1^ images acquired using the Hyperfine scanner [14] at the CURE Children’s Hospital in Uganda. We consider two important clinical questions: a Two Class Problem (PIH and NPIH with paired FLAIR and CT) and a Three Class Problem (PIH, NPIH, and healthy samples with FLAIR). GLAPAL-H outperformed state-of-the-art methods, including BAR-Net with high-quality CT scans, across various metrics. It demonstrated robust performance in infection detection, even with dataset perturbations and reduced training data. Ablation studies and activation maps highlighted the complementary nature of global and local branches, with the local branch showing superior individual performance for infection detection. GLAPAL-H offered enhanced interpretability, revealing detailed infection-activating regions. These results indicate GLAPAL-H’s potential to advance automated diagnosis of pediatric hydrocephalus in resource-constrained settings, effectively utilizing low-field MRI and incorporating domain-specific knowledge for improved diagnostic accuracy and efficiency in clinical practice.

## II. Methods: GLAPAL-H (Global, Local, And Parts Aware Learner for Hydrocephalus)

This project was performed on deidentified human image data following informed consent under the oversight of the Institutional Review Boards of Yale University, Penn State University, and the Research Ethics Committee of the Mbarara University of Science and Technology. The CT scans were deidentified before sharing with researchers. Low-field MRI scans were processed in two data streams – one with full personal health information directed to the local server at the point-of-care for clinical use, and another fully deidentified stream to the Hyperfine Cloud, where researchers were able to access these data under agreement with Hyperfine and with IRB approval. For infant studies, IRB approval is also obtained to share birth dates and dates of scans to characterize images at young ages.

### A Notation

We begin by introducing the necessary notation. Given *N* 2D image slices *S*_1_, *S*_2_, … , *S*_*N*_ for an infant, we compute the logit score for each slice *S*_*i*_ and ensemble the scores to obtain the final score, as described in Section II-E. Let *X* ∈ R^*H×W*^ denote the image (height *H*, width *W* ), obtained after performing the data preprocessing (Section III-A) on the FLAIR / CT scan. The variable *Y* denotes the corresponding infant’s label, indicating either NPIH, PIH, or Healthy. The input tensor *X* is processed by the network *f* (*·* , *θ*), yielding the output logit *Ŷ* = *f* (*X, θ*), where *θ* denotes the vector of all parameters within the neural network. Furthermore, we have a manually labeled (by medical professional) segmentation map *M* , which divides the image into three regions - brain tissue, CSF, and non-brain region respectively, relative to *X*. The network *f* (*·, θ*) comprises three branches: the segmentation branch, the global branch, and the local branch, each elaborated in Section II-C. The loss function *L*(*θ*) is minimized to yield 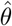 and a respective reasonable estimate of *f* ( *·* , *θ*) as 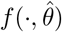. *L*(*θ*) and the training strategy are described in detail in Sections II-D and III-B respectively. Section II-B discusses the rationale behind the novel design of GLAPAL-H.

### B. Rationale behind GLAPAL-H

Fig. 1 (D) from [5] shows empirically that the occurrence of septation, abscess, and debris of the brain (CT Score) is higher in PIH infants than in NPIH infants. Also, Fig. 1 (B-C) shows the class activation maps corresponding to the detection of PIH or NPIH by BAR-Net [39] with FLAIR images. The class activation maps show two distinct behaviors - Fig. 1(B) shows the activation maps, where the region of importance for class-specific activation is spread globally all over the brain. In contrast, Fig. 1(C) shows concentrated areas of activation in specific local regions. This depicts the importance of considering two kinds of feature extractors: one feature extractor should focus on finding the whole brain’s global features, and another feature extractor that concentrates on local regions of the brain image and looks for specific signs of infections. Moreover, as Fig. 1(D) from [5] suggests we have different visible features of infections inside the brain tissue and inside the CSF separately. Motivated by the rationale of hydrocephalic etiology manifestation in diagnostic imagery, we design the GLAPAL-H architecture to guide the model in a better way for infection diagnosis and detection, GLAPAL-H consists of three fundamental branches - global, segmentation, and local branches. The goal of the global branch is to find global-type features in a larger receptive field of the image. The segmentation branch’s goal is to learn important features for segmenting the brain into the brain tissue and CSF. The local branch’s goal is to extract local-type features from smaller receptive sections of the image along with guidance by segmentation of the brain and CSF using the segmentation branch. The local branch builds a relatively less deep neural network to make sure that the model looks at local regions instead of larger global regions since the smaller depth of a convolutional neural network implies a smaller aggregate receptive field. To make sure that the global and local features are learned in the global and local branches in a complementary way, three unique regularizers are defined (in Section II-D) to help the architecture realize its goals.

### C GLAPAL-H Architecture

The proposed architecture *f* ( *·* , *θ*) as shown in Fig. 2. comprises three branches: a segmentation branch (red), a global branch (yellow), and a local branch (green). The segmentation branch aims to learn features for accurate segmentation, which serve as input features for the local branch to generate learnable parts masks for local feature extraction in brain tissue, and CSF. The global and local branches are designed to capture global and local features, respectively, for enhanced hydrocephalic infection diagnosis, as motivated in Section II-B. GLAPAL-H integrates the complementary information from these branches by fusing the outputs of the global and local branches and passing it through a linear layer *H* in the fusion head (blue) to obtain the logit (score) *Ŷ* for each input *X*.

**Fig. 2.**
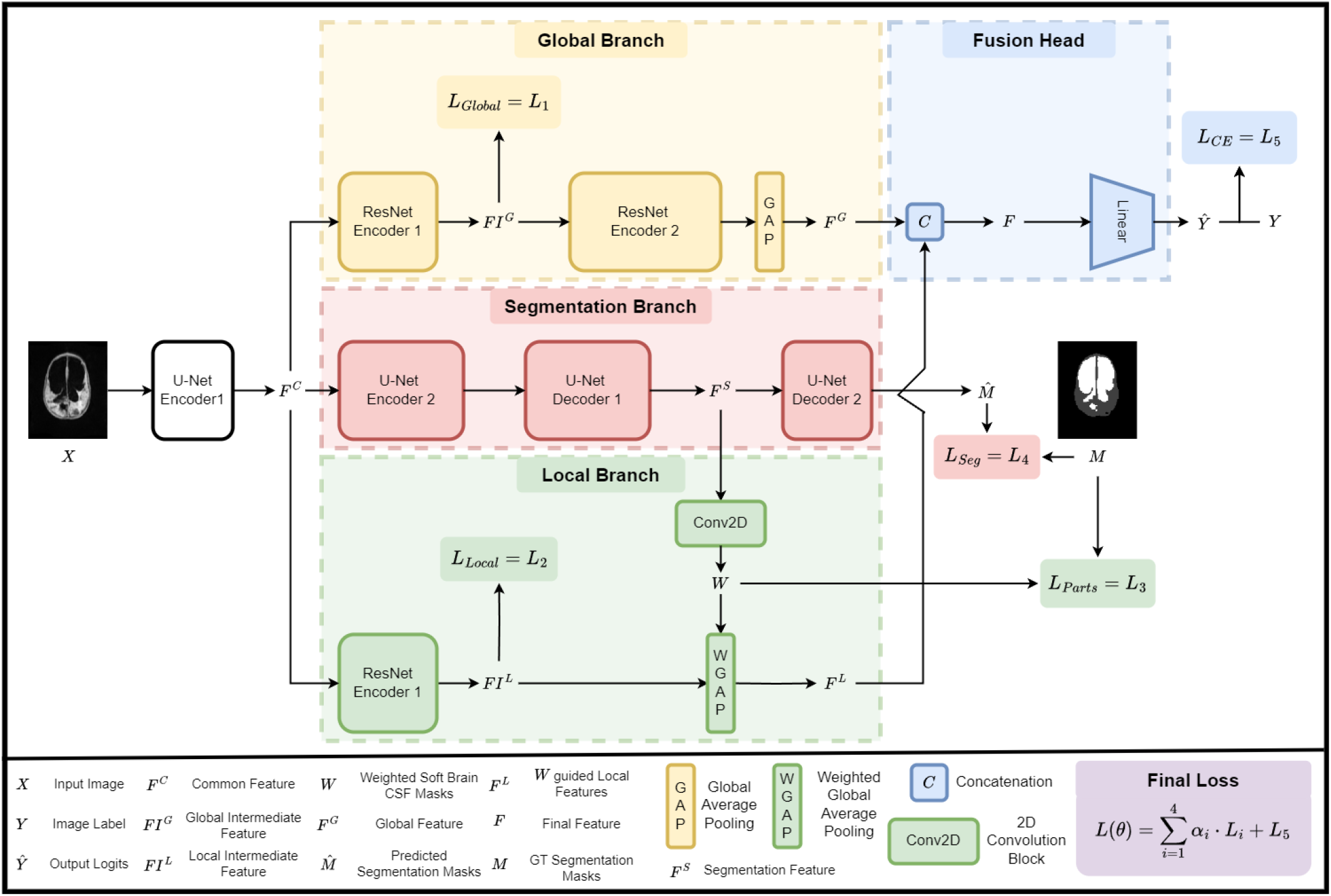
An overview of the GLAPAL-H model with three parallel branches: (1) the global branch for global feature extraction, (2) the segmentation branch, which employs brain tissue, and CSF mask prediction, and (3) the local branch for extracting the local features with parts-based brain tissue and CSF guidance. Features from the global and local branches are combined through a fusion head using concatenation and linear layers to get the classification outputs. The network optimizes a composite loss function, which includes contributions from the global, local, parts-based, segmentation, and cross-entropy losses.

#### 1) Segmentation branch

The segmentation branch in Fig. 2 consists of two encoders (U-Net Encoder 1, 2) followed by two decoders (U-Net Decoder 1, 2) with skip connections. Skip connections facilitate direct information flow between encoder and decoder layers, preserving spatial details for better feature reconstruction. The encoder and decoder together form the U-Net [65] architecture. The U-Net Encoder 1 consists of the first two convolutional layers, and the U-Net Encoder 2 comprises the remaining encoder layers of the U-Net. The U-Net Decoder 1 consists of the first ten convolutional layers of the U-Net decoder, while the U-Net Decoder 2 contains the remaining decoder layers predicting the segmentation masks 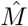 The outputs of the U-Net Encoder 1 and U-Net Decoder 1 are denoted as *F* ^*C*^ (common feature) and *F* ^*S*^ (segmentation feature), respectively. 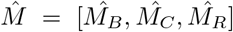 here 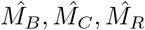 represent the predicted logits of the masks corresponding to brain tissue (B), CSF (C), and non-brain region (R), respectively.

#### 2) Global branch

The output of the U-Net Encoder 1 block, *F* ^*C*^, is taken as an input to the global branch, a truncated version of the pre-trained ResNet-18 [66] adjusted to the input dimensions of *F* ^*C*^. Since the U-Net Encoder 1 consists of the first two convolutional layers, *F* ^*C*^ has 64 as the channel dimension. Hence, the first convolutional and pooling layers of ResNet-18 are removed, and the rest of the architecture is retained up to the Global Average Pooling layer [66]. This step utilizes the pre-trained ResNet-18 model on ImageNet [66] and truncates it to take input with 64 channels. The truncated ResNet-18 now has 16 convolutional layers with residual connections. The global branch is divided into the ResNet Encoder 1 and the ResNet Encoder 2, followed by the Global Average Pooling layer. The ResNet Encoder 1 consists of the first four convolutional layers with residual connections, while the ResNet Encoder 2 comprises the remaining 12 convolutional layers. With the input *F* ^*C*^, the ResNet Encoder 1 produces an output *FI*^*G*^ (intermediate global feature), which is then passed through the ResNet Encoder 2, along with the Global Average Pooling (GAP) layer, to generate an output *F* ^*G*^ (global feature) with 512 channels. We incorporate the global regularizer *L*_*Global*_ on *FI*^*G*^ to extract global features, complementing the local features (introduced in Section II-D).

#### 3) Local branch

The local branch consists of an encoder ResNet Encoder 1 followed by a Weighted Global Average Pooling (WGAP) layer guided by the segmentation features *F* ^*S*^. The output from the U-Net Encoder 1 block: *F* ^*C*^ is taken as input to the encoder ResNet Encoder 1, which is architecturally similar to the ResNet Encoder 1 block in the global branch. Note that the ResNet Encoder 1 blocks in the global and local branches are distinct encoders with no shared weights. The output of the ResNet Encoder 1 block is *FI*^*L*^ (intermediate local feature), which has a constraint as a local regularizer *L*_*Local*_ to learn the local features (described in Section II-D). The output from the U-Net Decoder 1 *F* ^*S*^ is passed through a learnable 2D Convolution layer to obtain *W* = [*W*_*B*_, *W*_*C*_] with two channels, representing learnable soft masks similar to the original segmentation masks. This step aims to create soft masks *W*_*B*_, *W*_*C*_ instead of hard binary segmentation masks, useful for capturing local features inside the brain tissue and CSF, respectively. This demands a regularizer for *W* , explained in Section II-D. *FI*^*L*^ has 64 channels, while each output channel of *FI*^*L*^, and *W*_*B*_, *W*_*C*_ have different feature dimensions. We make sure that each feature output of *FI*^*L*^, and *W*_*B*_, *W*_*C*_ are of the same size by using nearest-neighbor interpolation on *W*_*B*_, *W*_*C*_. We perform a Hadamard product of *FI*^*L*^ with *W*_*B*_, *W*_*C*_ individually, followed by a Global Average Pooling (GAP) to obtain two features *F*_*B*_, *F*_*C*_, where *F*_*B*_ = *W*_*B*_ *·FI*^*L*^, and *F*_*C*_ = *W*_*C*_ *· FI*^*L*^. Each of *F*_*B*_, *· F*_*C*_ has 64 channels, which we concatenated to get *F* ^*L*^ = [*F*_*B*_, *F*_*C*_] with 128 channels. The layer that transforms *FI*^*L*^ into *F* ^*L*^ using the weighted masks *W* is named as Weighted Global Average Pooling (WGAP) layer. The complementarity of the local branch and the global branch is noteworthy since the local branch ensures that the collected features are local, as the number of layers in the local branch is much less than in the global branch. The number of layers makes sure that the receptive field of the local branch is much smaller than the global branch, which leads to the extraction of localized features, which are guided by the parts-based guidance.

#### 4) Fusion head

The features from the global and local branches, *F* ^*G*^ and *F* ^*L*^, are concatenated to create a feature *F* with dimension 640, followed by a fully connected layer head *H* to obtain the logits *Ŷ* for the classification classes. Section II-D introduces the cross-entropy loss based on *Ŷ* and the ground truth labels *Y* . In the following sections, we introduce different regularizers and loss functions: one from the global branch, one from the segmentation branch, two from the local branch, and one from the final fusion head.

### D. GLAPAL-H Loss Functions

The proposed GLAPAL-H architecture incorporates five regularizers and loss functions: *L*_*Global*_, *L*_*Local*_, *L*_*P arts*_, *L*_*Seg*_, and *L*_*CE*_ (cross-entropy loss), which are represented in Fig. 2 as *L*_1_, *L*_2_, *L*_3_, *L*_4_, and *L*_5_ respectively. These components are designed to ensure effective learning of complementary global and local features, accurate segmentation, and robust classification performance. The local and parts regularizers of the local branch and the global regularizer of the global branch are introduced to make sure that the model learns the global features and not the detailed local features, which the local branch will handle. This careful engineering is important to craft a complementary relationship between the global and local branches. To define the regularizers of each feature, we will assume that each of the features is of the dimension *C × H × W* for mathematical brevity. We also name each of the individual regularizers using *L*_*i*_ to accommodate space for defining final loss function *L*(*θ*).

#### 1) Global Regularizer - *L*_*Global*_

The global regularizer *L*_*Global*_ is introduced to capture global features during the learning process. To achieve this, a feature variation loss is proposed on *FI*^*G*^. A feature variation loss *FV L*(*χ*) is defined on an intermediate feature *χ* with dimension *C ×H × W* , *N* = *CHW* , and 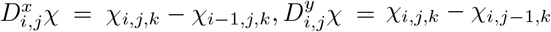 as

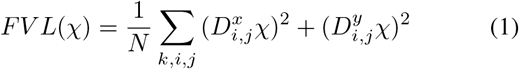

The feature variation loss on the intermediate global feature *FI*^*G*^, that defines our global regularizer,

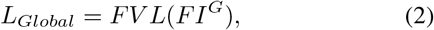

aims to capture low-level features, such as the shape and structure of the entire brain region, and send it to the next layers for global feature extraction like large-scale global asymmetry. This loss is inspired by the total variation norm [67]. In past work, the total variation loss has been used for image denoising and restoration that achieves edge preservation in the presence of noise. It has been shown to smooth out images while preserving core structural elements. By forcing the minimization of a variation loss on the extracted features, we are essentially encouraging the features to be representative of the global image content, leading to a complementary collaboration of the global and local branches.

#### 2) Local Regularizer - *L*_*Local*_

The local regularizer *L*_*Local*_ is introduced to capture local features during the learning process. To achieve this, a contrast regularizer is proposed on *FI*^*L*^. A contrast regularizer *CR*(*χ*) is defined on an intermediate feature *χ* with dimension *C × H × W* , *N* = *CHW* , and 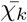is the average image for the *k*^*th*^ channel, where 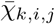 denotes the (*i, j*)^*th*^ pixel of the average 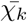 image. The contrast regularizer is defined as

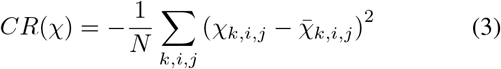

The contrast regularizer aims to capture high-level detailed features, such as the signs of infections in the local regions by optimally increasing the contrast of *FI*^*L*^:

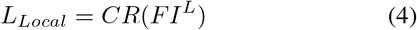

The regularizer is applied to *FI*^*L*^, the feature output of a significantly shallower network compared to the global branch. As a result, the local branch has a much smaller receptive field, facilitating the extraction of precise localized features.

#### 3) Parts regularizer - *L*_*P arts*_

The segmentation mask *M* is transformed into individual binary masks *M*_*B*_, *M*_*C*_, *M*_*R*_ corresponding to 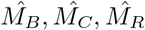. The parts regularizer is defined to ensure that *W*_*B*_ and *W*_*C*_ are softer versions of the hard masks *M*_*C*_ and *M*_*B*_, respectively, which learn features within the segmentation region but for classification. While the traditional segmentation masks typically focus on only brain tissue and CSF for segmentation, the softer version of *M* , denoted as *W* , not only leverages segmentation for guidance but also enhances classification performance, by enabling more efficient distribution of weights within the brain tissue and CSF, resulting in improved discriminative features for infection diagnosis within the brain tissue and CSF parts. The 2D convolution layer which uses the segmentation features to output *W* learns the necessary features to distribute the weights for infection diagnosis, which preserves the information on the segmentation of the brain tissue, and CSF.

The dice loss between two-dimensional images *A* and *B* with image pixel *A*_*i*,*j*_, and *B*_*i*,*j*_ respectively (with *ϵ* is a small positive constant added for numerical stability) is defined as:

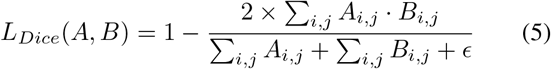

We define the parts regularizer as follows:

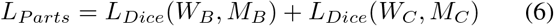

where *W*_*C*_ and *W*_*B*_ are the soft masks, *M*_*C*_ and *M*_*B*_ are the corresponding hard masks. Also, the *M*_*C*_ and *M*_*B*_ are downscaled versions to match the size of *W*_*C*_ and *W*_*B*_. The dice loss computes the similarity between the soft masks (*W*_*C*_ and *W*_*B*_) and the hard masks (*M*_*C*_ and *M*_*B*_). The loss is minimized when the soft masks closely approximate the hard masks, ensuring that the local branch learns local features, and is guided by the segmentation into brain tissue, and CSF.

#### 4) Segmentation loss - *L*_*Seg*_

The segmentation loss uses dice loss to ensure efficient learning of the segmentation features *F* ^*S*^. The segmentation loss *L*_*Seg*_ is defined as:

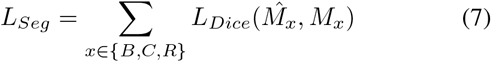

#### 5) Cross Entropy Loss - *L*_*CE*_

The *L*_*CE*_, based on the classification labels is the standard cross-entropy loss between *Ŷ* and *Y* , defined as follows for a batch size *b*, and *k* classes:

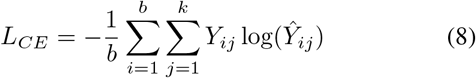

Here, *Y*_*ij*_ = 1 if the *i*^*th*^ sample belongs to the *j*^*th*^ class, and 0 otherwise, and *Ŷ*_*ij*_ represents the predicted logit that the *i*^*th*^ sample belongs to the *j*^*th*^ class.

#### 6) Final Loss - *L*(*θ*)

The final loss *L*(*θ*) is defined as a linear combination of the five regul defined above as 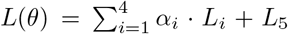 where *α* = (*α*_1_, *α*_2_, *α*_3_, *α*_4_) is a tuple of hyperparameters, which are to be determined by cross-validation. The specific value of *α* is in Section III B.

### E. Ensemble of Predictions

GLAPAL-H predicts a logit (score) for each of the *N* slices of an infant. To compute the final logit for an infant, only the middle 50% of slices are considered. Let the predicted logits for the *N* slices be *Ŷ*_1_, *Ŷ*_2_, … , *Ŷ*_*N*_ . The final predicted logit for the infant is calculated as:

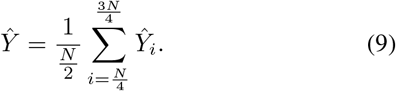

This method ensures that the final logits are derived from the central slices, which are likely to contain more representative information while reducing the influence of the edge slices, seriously affected by the movement of the infants (since they were not sedated). The final predicted label for the infant is determined as arg max_*j*_ *Ŷ*_*j*_ , where *j* represents the index of the class, and *Ŷ*_*j*_ is the predicted logit for class *j* (after softmax). This ensures that the predicted label corresponds to the class with the highest logit in the final aggregated logits *Ŷ* .

## III. Experiments and Results

### A. Dataset and Data Preprocessing

The dataset for this study was collected at CURE Children’s Hospital of Uganda (CCHU), a nationwide neurosurgical referral center located in Mbale, in the eastern region of Uganda. A total of 73 infants were included in the study. The dataset comprises 45 infants with paired low-field Fluid Attenuated Inversion Recovery (FLAIR) MRI and Computed Tomography (CT) images: 26 were labeled as Post-Infectious Hydrocephalus (PIH), 19 were labeled as Non-Post-Infectious Hydrocephalus (NPIH), and 28 infants with FLAIR images were labeled as Healthy. Each infant’s FLAIR sequence contains approximately 40 slices, while there are around 35 for each CT scan sequence. The classification of PIH and NPIH was based on established clinical criteria. It is important to note that CT scans were not performed on healthy infants due to the increased cancer risk associated with radiation exposure. As a result, the healthy cohort only includes FLAIR images. For the FLAIR images, we also have ground truth segmentation masks annotated by medical professionals using ITK-Snap [68], which are divided into three classes: brain tissue, cerebrospinal fluid (CSF), and non-brain region. All the scanned images were collected in DICOM format. The FLAIR and CT images were skull-stripped using image processing algorithms similar to the method mentioned in [39] to remove background information. The CT images in DICOM format were transformed by an affine transformation using the rescale and intercept values in the DICOM header data of the corresponding slice sequences of individual infants for proper contrast adjustment before the skull strip. The FLAIR images didn’t undergo any such transformation before the skull strip. **Two and Three Class Problems:** Two and Three Class Problems are important medical questions for differentiating healthy and hydrocephalus infants and diagnosing infection among patients. We conducted extensive experiments for both of these problems and examined the performance of GLAPAL- H compared to state-of-the-art alternatives. Labeled images are used from the aforementioned dataset to investigate - **Two Class Problem**, which includes two classes - Post-Infectious Hydrocephalus (PIH) and Non-Post-Infectious Hydrocephalus (NPIH) with paired FLAIR and CT for 47 infants, and **Three Class Problem**, which includes three classes - PIH, NPIH, and Healthy for 73 infants with only FLAIR imagery.

**TABLE I.** Comparison of State of the Art Methods for PIH vs. NPIH (Two Class) problem.

| Model | Accuracy (Acc) | Sensitivity | Specificity | F1 Score | AUC | P-value (Acc) |
| --- | --- | --- | --- | --- | --- | --- |
| **GLAPAL-H** | **87.11** | **92.67** | **80.00** | **89.04** | **87.89** | - |
| BAR-Net CT [39] | 84.61 | 88.28 | 78.34 | 86.20 | 80.06 | <0.05 |
| BAR-Net [39] | 82.67 | 88.67 | 73.33 | 85.92 | 77.56 | <0.01 |
| MedNeXt [69] | 80.89 | 92.01 | 63.72 | 85.24 | 83.78 | <0.01 |
| MedViT [70] | 79.11 | 89.27 | 63.15 | 83.33 | 81.61 | <0.01 |
| RS<sup>2</sup>Net [48] | 70.22 | 88.29 | 42.10 | 78.12 | 68.39 | <0.01 |

**TABLE II.** Comparison of State of the Art for Healthy vs. PIH vs. PIH (Three Class) Problem.

| Model | Accuracy (Acc) | Sensitivity | Specificity | F1 Score | AUC | P-value (Acc) |
| --- | --- | --- | --- | --- | --- | --- |
| **GLAPAL-H** | **90.19** | **87.89** | **94.91** | **88.25** | **91.79** | - |
| MedNeXt [69] | 87.71 | 85.44 | 92.72 | 85.80 | 89.45 | <0.05 |
| BAR-Net [39] | 86.29 | 83.78 | 91.89 | 84.03 | 87.57 | <0.05 |
| MedViT [70] | 84.81 | 81.61 | 91.06 | 81.97 | 88.58 | <0.01 |
| RS<sup>2</sup>Net [48] | 80.67 | 78.89 | 86.44 | 80.16 | 85.68 | <0.01 |

### B Experimental Setup

#### 1) Implementation Settings and Training Strategy

Five-fold nested cross-validation was employed for both the Two Class and Three Class classification tasks. In each fold, 80% of patients and their corresponding scans were used for training, while the remaining 20% of patients and scans were set aside for testing. Stratified sampling ensured that the class distribution in each fold mirrored that of the original dataset. A model generates a classification label based on the ensemble of predicted scores for the slices as discussed in section II-E. Each of the scans and segmentation masks of size (*H, W* ) was transformed into a square image of size (*M, M* ), where *M* = max *{H, W}* for rotation augmentation without information loss. This step was important because, due to the movement of the infants, the image centers had shifted to distinct positions. Several data augmentation operations were employed jointly on each scan and its segmentation mask, including flipping, randomly rotating − 20^*°*^ to 20^*°*^ in the axial plane, cropping, and resizing. The model was optimized using the Adam optimizer [71] with a mini-batch size of 32. To reduce computational load, GLAPAL-H was trained in the following way. The segmentation branch was trained and saved initially for each fold. During the GLAPAL-H’s global and local branches’ training for the training data, the entire model was trained again for each fold. The outputs *F* ^*C*^ and *F* ^*S*^ were extracted from the saved segmentation branch for each validation fold. During validation or testing, no original segmentation masks were used. This training strategy is equivalent to minimizing *L*_*Seg*_ = *L*_4_ first, and then minimizing the rest of the loss function 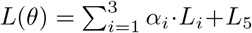. Each model was trained for 100 epochs with early stopping. The initial learning rate was set to 0.0001 and was multiplied by a factor of 0.1 after every 10 epochs. The hyperparameter values (*α*_1_, *α*_2_, *α*_3_) were selected to be (0.01, 0.02, 0.01). A weight decay of 0.0001 was used. The optimal hyperparameters were chosen by further dividing the training examples into 80% training and 20% validation data, making the overall training-testing strategy similar to nested five-fold cross-validation [72]. We verified that GLAPAL-H’s performance was robust in the vicinity of the aforementioned optimal hyperparameter values as confirmed experimentally in Fig. S-1 of the Supplementary document. The training, validation, and evaluation were carried out using the Pytorch framework [73] on an NVIDIA TITAN RTX GPU with 24 GB of memory.

#### 2) Evaluation Metrics

Performance was assessed using accuracy, sensitivity, specificity, F1 score, and area under the ROC curve (AUC). We employed macro-averaging-based metrics [74] for the multi-class problem. Statistical significance was assessed using appropriate methods [75] with a significance level of 0.05. Confusion matrices are also reported, where each cell value is normalized as a percentage relative to the true label occurrences. Confusion matrices help evaluate class-wise performance by showing how well each class is correctly predicted and where misclassifications occur, enabling a robustness analysis of the model’s performance.

In the upcoming sections, we analyze the effectiveness of the local and global branches and the parts-based guidance on GLAPAL-H’s performance in the Ablation Study in the Two Class Problem. Then, we compare GLAPAL-H with the state-of-the-art methods for both the Two Class and Three Class problems. Finally, we compare the interpretability of GLAPAL-H’s local and global branches along with the overall class activation map (CAM) for infection diagnosis with the top two performing models for the Two Class Problem.

### C. Ablation Study

We conducted an ablation study to evaluate the impact of the architectural branches and the corresponding regularizers on GLAPAL-H’s performance in the Two Class Problem. Table III presents the results of this analysis. The columns “*L*_*Global*_”, “*L*_*Local*_”, and “*L*_*P arts*_” indicate the presence or absence of regularizers in the respective components. **1) Baseline Performance:** When all regularizers are absent, GLAPAL-H achieves a relatively lower accuracy. **2) Synergistic Effect:** The combination of global and local branches with regularizers without the parts regularizer yields a slight improvement in accuracy, demonstrating the complementary nature of global and local branches, and their features as extracted by the *L*_*Global*_ and *L*_*Local*_ regularizers. The two branches, along with their regularizers, should work together for an ideal performance. 3)**Parts Regularizer Contribution:** The inclusion of the parts regularizer along with global and local regularizers and architecture provides the highest accuracy, underscoring the importance of part-based segmentation-guided feature extraction inside brain tissue and CSF, respectively, for infection diagnosis and hydrocephalus detection. The ablation results highlight the critical role of local and part-based features in the task at hand, with the original GLAPAL-H architecture incorporating all three components providing the most effective configuration. The significant performance boost observed when including the parts regularizer suggests that fine-grained, part-based analysis of brain structures is particularly valuable for this medical imaging task, aligning with clinical intuition that subtle changes in specific brain regions are crucial for accurate diagnosis of hydrocephalus infection detection.

**TABLE 3.** Ablation study of the effect of local and global branches on the mean accuracy for the Two Class Problem.

| $L_{Global}$ | $L_{Local}$ | $L_{Parts}$ | Accuracy |
| --- | --- | --- | --- |
| Yes | Yes | Yes | 87.11 |
| Yes | Yes | No | 84.68 |
| No | No | No | 83.33 |

### D Comparisons against SOTA methods

To assess the effectiveness of our proposed method, GLAPAL-H, we compare it against state-of-the-art (SOTA) approaches in medical image classification. GLAPAL-H leverages hydrocephalus etiology-guided global and parts-based local feature extraction to effectively process low-quality images. The selected SOTA methods reflect the progression of computer vision techniques in medical imaging from 2012 to the present: **BAR-Net** [39], a CNN-based model that highlights the dominance of convolutional architectures in medical image classification during the 2012–2019 period, demonstrating the capability of CNNs to extract meaningful features from medical images; **MedViT** [70], a hybrid CNN-Vision Transformer (CNN-ViT) model representing the transition period (2019–2021) when Vision Transformers (ViTs) began emerging as a viable alternative in medical imaging tasks, combining the strengths of both CNNs and ViTs; **MedNeXt** [69], based on the ConvNeXt architecture, exemplifies the latest advancements in CNN design, offering improved performance and efficiency over traditional CNNs; and **RS**^2^**Net** [48], a segmentation-guided classification method that incorporates segmentation information into the decision-making process.

It’s important to note that all the aforementioned SOTA methods were primarily developed and tested on highresolution medical imagery, such as CT, MRI, X-ray, or Ultrasound scans. This context is crucial when interpreting their performance relative to our proposed method. By benchmarking against these diverse SOTA methods, we provide a comprehensive evaluation of GLAPAL-H’s effectiveness across different architectures and methodologies in medical image classification. We first evaluate the performance of GLAPAL-H against SOTA alternatives for the classification task in the Two Class Problem using the evaluation metrics.

#### 1 Two Class Problem

Table I presents the mean and standard deviation of performance measures for each method, while Fig. 3 illustrates the Receiver Operating Characteristic (ROC) curves. The corresponding Area Under the Curve (AUC) values are reported in the last column of Table I. Our comprehensive analysis reveals that GLAPAL-H consistently outperforms state-of-the-art (SOTA) methods across all key performance metrics. GLAPAL-H demonstrates superior accuracy, sensitivity, and specificity compared to BAR-Net, MedViT, MedNeXt, and R*S*^2^Net. The AUC values further corroborate GLAPAL-H’s enhanced discriminative capability in the classification task. Notably, RS^2^Net, which utilizes segmentation information, shows the lowest performance among the compared methods. This suggests that our approach of incorporating local and global features is more effective than this particular segmentation-guided method for the given task. GLAPAL-H’s superior performance can be attributed to its domain-enriched local and global architecture with regularizers specifically designed for hydrocephalus classification.

**Fig. 3.**
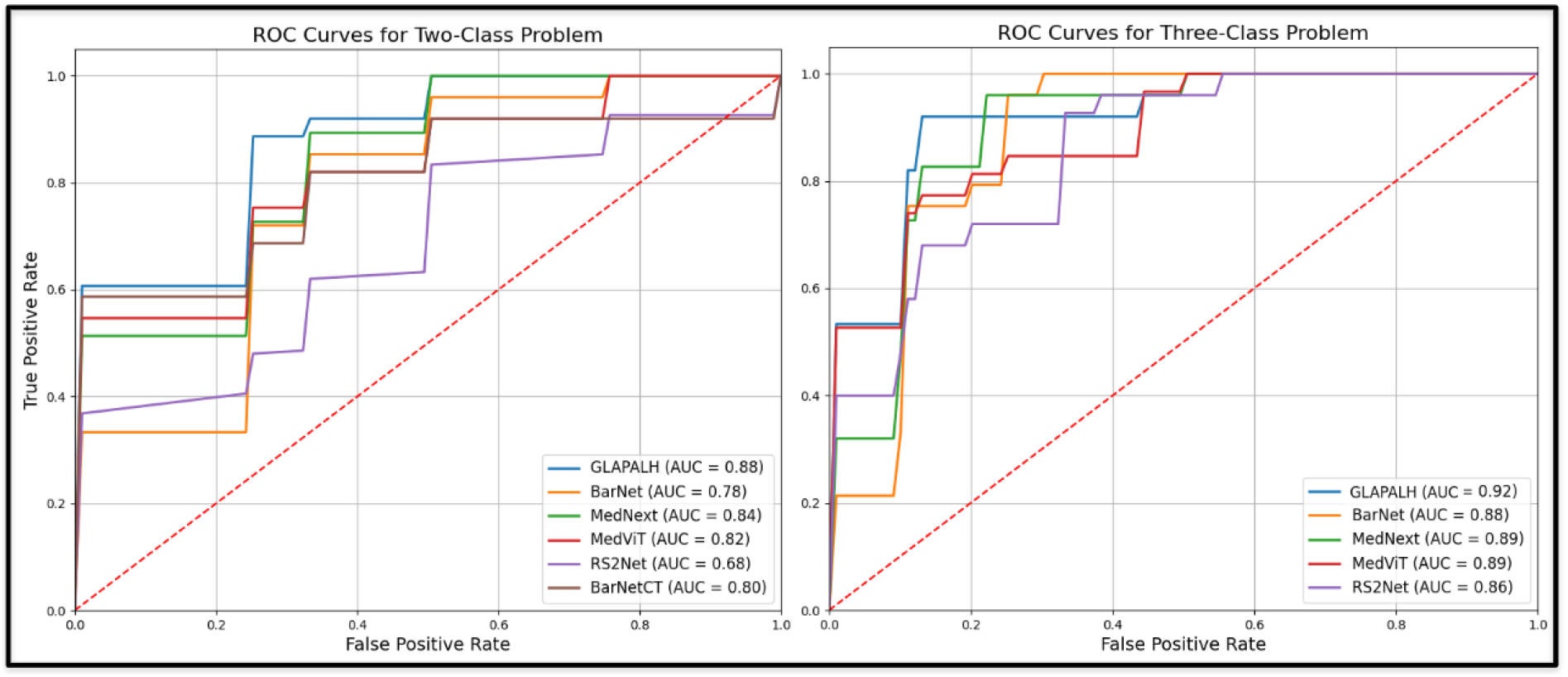
ROC Curves of GLAPAL-H and SOTA methods for both the Two Class and the Three Class Problems.

#### 2) Three Class Problem

The Three Class Problem represents a clinically important case where healthy infants must be separated simultaneously from hydrocephalic infants, and results for this setting are summarized in Table II. Overall, the metrics of Table II show how GLAPAL-H performs better than all the SOTA methods. The confusion matrices in Fig. 4 for the top three methods (GLAPAL-H, MedNeXt, and BAR-Net) reveal perfect classification of healthy patients, with no false healthy signals. However, the distinct visual information and intensity distribution in healthy patients’ brain imagery can potentially confuse models and reduce their ability to differentiate between infection and non-infection cases. The individual detection rates for NPIH and PIH show interesting variations: GLAPAL-H’s NPIH detection decreased from the Two Class to the Three Class Problem, while PIH detection remained stable; BAR-Net’s PIH and NPIH detection rates both decreased significantly, which is expected given its specific design for Hydrocephalus PIH vs NPIH detection; and MedNeXt’s PIH detection rate decreased, while NPIH detection increased slightly, though still lower than GLAPAL-H’s performance. While overall accuracy increased for all models, this improvement is primarily due to accurate healthy patient classification, with the introduction of different data distributions (Healthy class) perturbing PIH and NPIH detection effectiveness in all top three models.

**Fig. 4.**
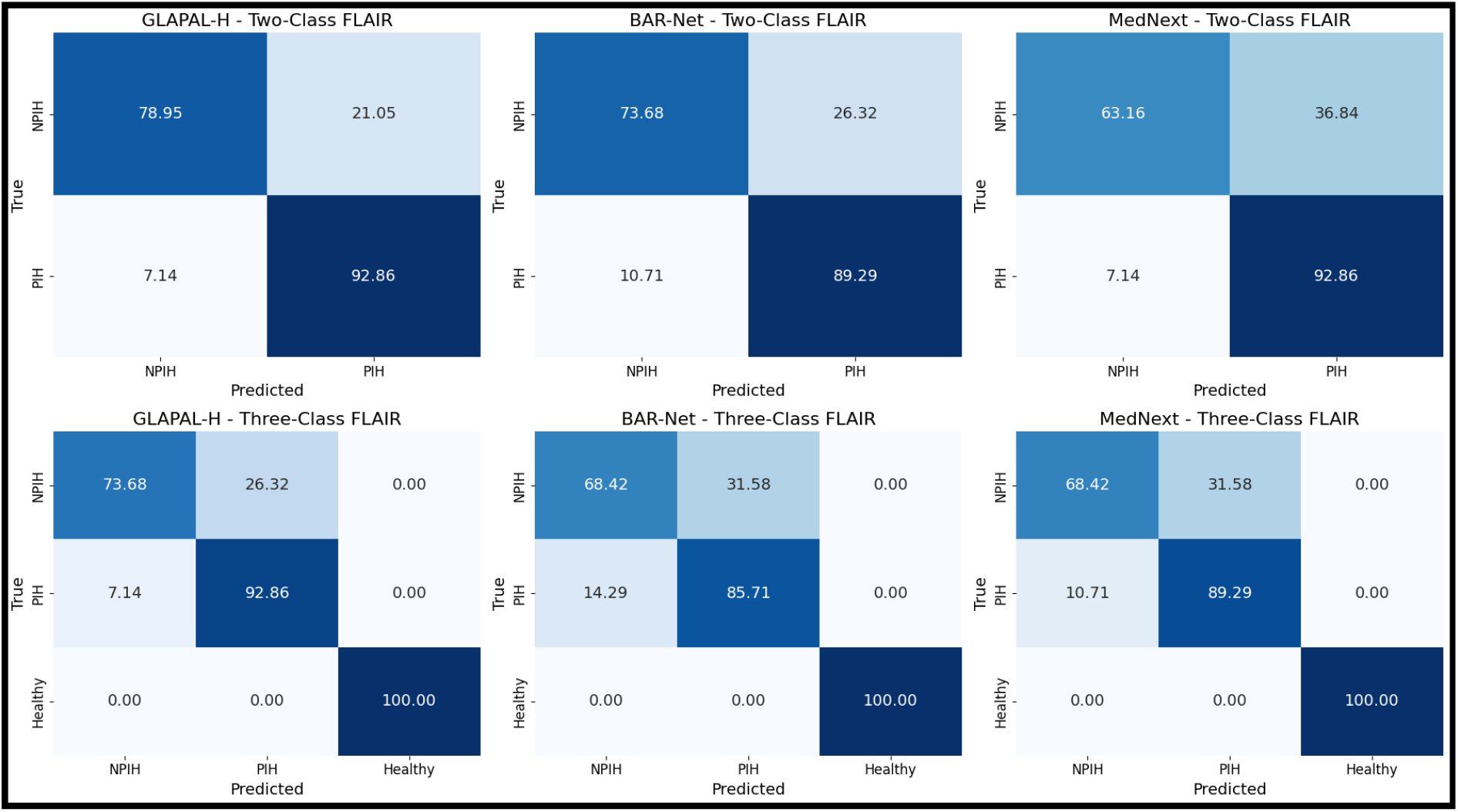
Normalized confusion matrices across labels for the Two Class and Three Class Problems of the top three methods: GLAPAL-H, BAR-Net, and MedNeXt on FLAIR: showing the superiority of GLAPAL-H in robust infection detection.

#### 3) Robustness

This scenario provides an opportunity to assess model robustness for infection diagnosis with a shift in training data distribution, with GLAPAL-H demonstrating superior robustness by maintaining consistent PIH (infection) detection rates and relatively smaller performance perturbation from the Two Class to the Three Class Problem, as illustrated in Fig. 5. This is interesting to see how the GLAPAL-H’s performance of infection detection remains consistent among both the Two Class and Three Class Problems, unlike the other methods MedNeXt and BAR-Net, which change the relative performance in two different problems. This gives evidence of the robustness of GLAPAL-H over SOTA alternatives.

**Fig. 5.**
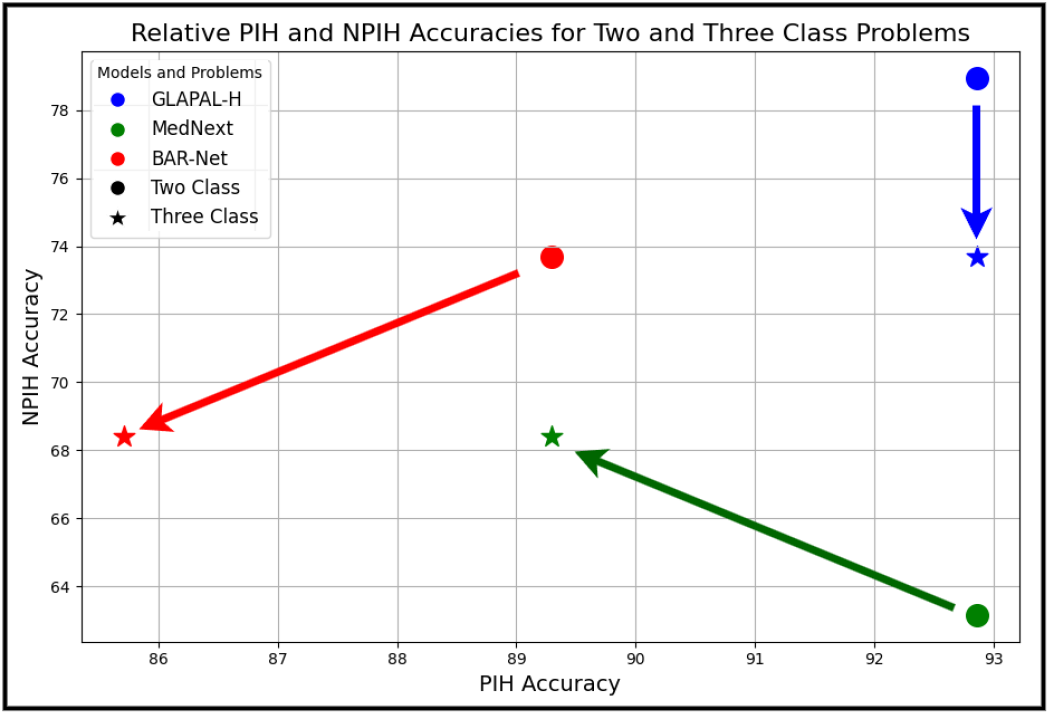
GLAPAL-H shows better robustness considering the perturbation of performance from the Two Class to the Three Class Problem when compared to MedNeXt and BAR-Net.

#### 4) Low Training

GLAPAL-H’s robustness is further demonstrated by its performance under low-training conditions in the Three Class Problem. Experiments conducted with only 40% of the original training dataset, while keeping the same validation and test datasets for each fold, revealed several key insights. As shown in Fig. 6, when the training data was reduced, MedNeXt experienced a performance drop twice as large as that of GLAPAL-H. Additionally, MedNeXt exhibited greater performance variability, as indicated by the error bars representing the standard deviation of accuracy, compared to GLAPAL-H. Notably, with just 40% of the training data, GLAPAL-H’s performance was nearly equivalent to that of MedNeXt using the full training set. These findings highlight GLAPAL-H’s robustness in data-visual cues for classification.

**Fig. 6.**
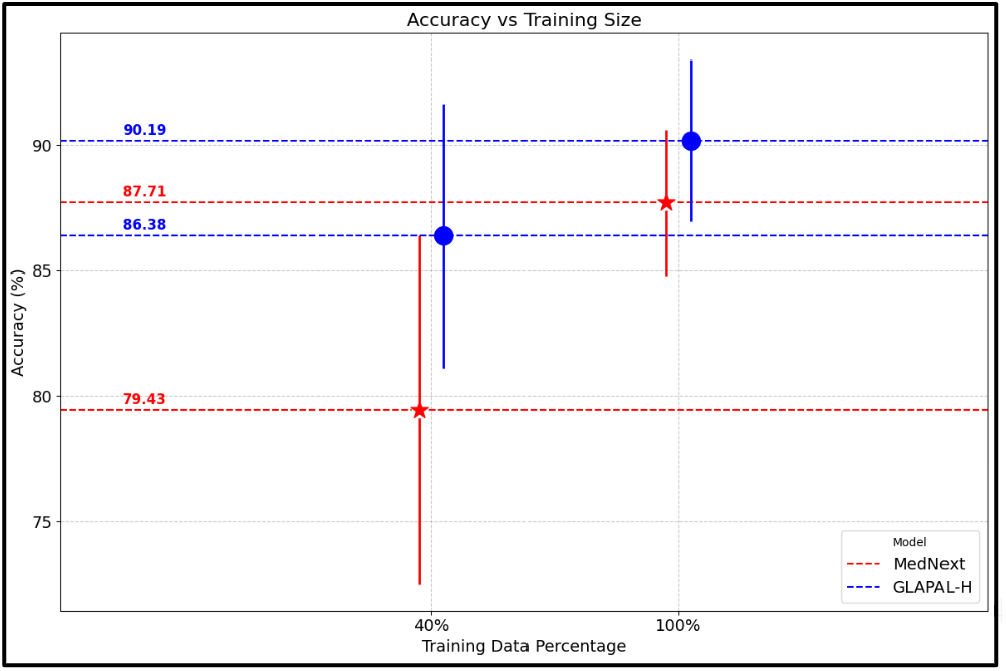
Low Training vs High Training Performance of GLAPAL-H and MedNeXt (Three Class Problem) with 40% and 100% of the training dataset, respectively.

#### 5 Computational Requirements

Table IV compares the computational complexity, training, and inference time across various models. Among the evaluated models, GLAPAL-H achieves a favorable trade-off between computational efficiency and performance. Although its parameter count is slightly higher than some comparators, GLAPAL-H exhibits reduced training times and comparable inference latency. This advantage is attributed to its disease etiology-guided learning framework, which strategically informs feature extraction and optimization, thereby promoting faster convergence. As a result, GLAPAL-H typically achieves early stopping after substantially fewer epochs compared to models lacking explicit clinical guidance. In contrast, deep architectures without *etiology guided prior knowledge* often require prolonged training and exhibit computationally burdensome convergence behavior, driven by reliance purely on available training. Notably, despite the inclusion of additional feature extraction branches, GLAPAL-H maintains inference times comparable to those of other state-of-the-art models. The incremental computational overhead is negligible relative to the observed gains in predictive performance. These characteristics position GLAPAL-H as a particularly promising solution for rapid, scalable deployment in resource-constrained clinical environments.

**TABLE 4.**
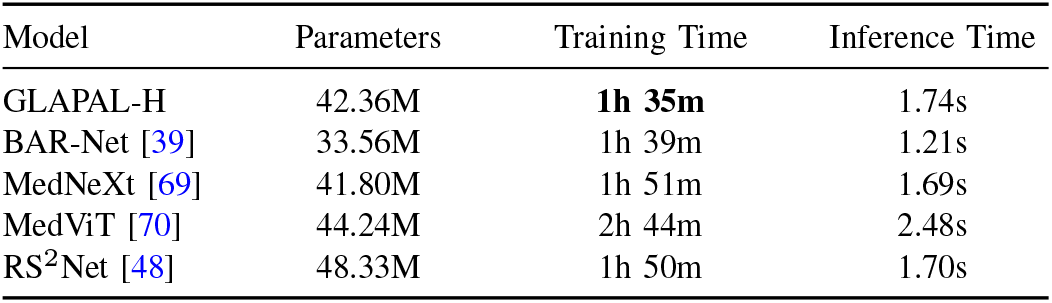
Comparison of Model Complexity, Training Time, and Inference Time (single infant) Two Class Problem.

| Model | Parameters | Training Time | Inference Time |
| --- | --- | --- | --- |
| GLAPAL-H | 42.36M | **1h 35m** | 1.74s |
| BAR-Net [39] | 33.56M | 1h 39m | 1.21s |
| MedNeXt [69] | 41.80M | 1h 51m | 1.69s |
| MedViT [70] | 44.24M | 2h 44m | 2.48s |
| RS<sup>2</sup>Net [48] | 48.33M | 1h 50m | 1.70s |

### E Activation Maps for Interpretability

Understanding the decisions made by deep learning models is essential for clinicians in treatment planning. As described in [76], a Class Activation Map (CAM) for image classification is calculated by taking a weighted sum of the feature maps, expressed as 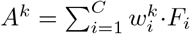. Here, *k* denotes the *k*^*th*^ class. *F*_*i*_ represents the *i*^*th*^ feature map from the final convolutional layer, which is defined below using the feature maps from the global and the local branches respectively. *C* is the total number of feature maps, and 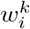 is the weight from the fully connected layer indicating the significance of the *i*^*th*^ feature map for class *k*. Regions with higher weights in *A*^*k*^ have a greater impact on predicting class *k*. To calculate the global activation map 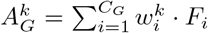 and local activation map 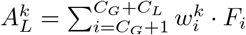, the feature maps are extracted from the final convolutional layer before features *F* ^*G*^, and *F* ^*L*^ from the global and the local branches, respectively. Since, the final linear layer maps *F* , which is the concatenation of *F*_*G*_ and *F*_*L*_ to the final logits, the weights are taken from the corresponding positions of *F*_*G*_ and *F*_*L*_ for the global and the local branches. We will now express the activation maps, mathematically. Let’s assume that *C* = *C*_*G*_ + *C*_*L*_, where *C*_*G*_ and *C*_*L*_ are the numbers of channels of the final convolutional layer in the global and the local branches. *F*_*i*_ for all 1 ≤ *i* ≤ *C*_*G*_ are the feature maps for the global branch, and *F*_*i*_ for all *C*_*G*_ + 1 ≤ *i* ≤ *C*_*G*_ + *C*_*L*_ are the feature maps for the local branch. All the activation maps are normalized between 0 and 1 for a better comparative study. Observe that 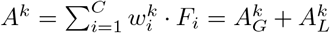. The impact of our unique architecture with innovative regularization in the global and local branches is demonstrated in the activation maps in Fig. 7 for two PIH patients. The complementarity of the local and the global branches is nicely captured in the local and the global branches. Observe how the global activation map captures the shape of overall CSF pockets along with debris, and the local activation maps capture the local signs of inflammation, necrosis, abscess, and loculation. This is remarkable when compared to the BAR-Net and MedNeXt, where the activation map spans a broad area without any specific region of importance for infection. Furthermore, the ablation study clearly demonstrates a more significant performance improvement from the local branch compared to the global branch, as evidenced by the local activation map’s successful detection of subtle, minute features. This provides interpretable evidence behind the superior performance of GLAPAL-H over the SOTA methods. For medical purposes, one should refer to the trinary combination of the activation maps of local, global, and the entire GLAPAL-H activation maps for a better understanding of the importance of various anatomical structures that may be visual cues for infection.

**Fig. 7.**
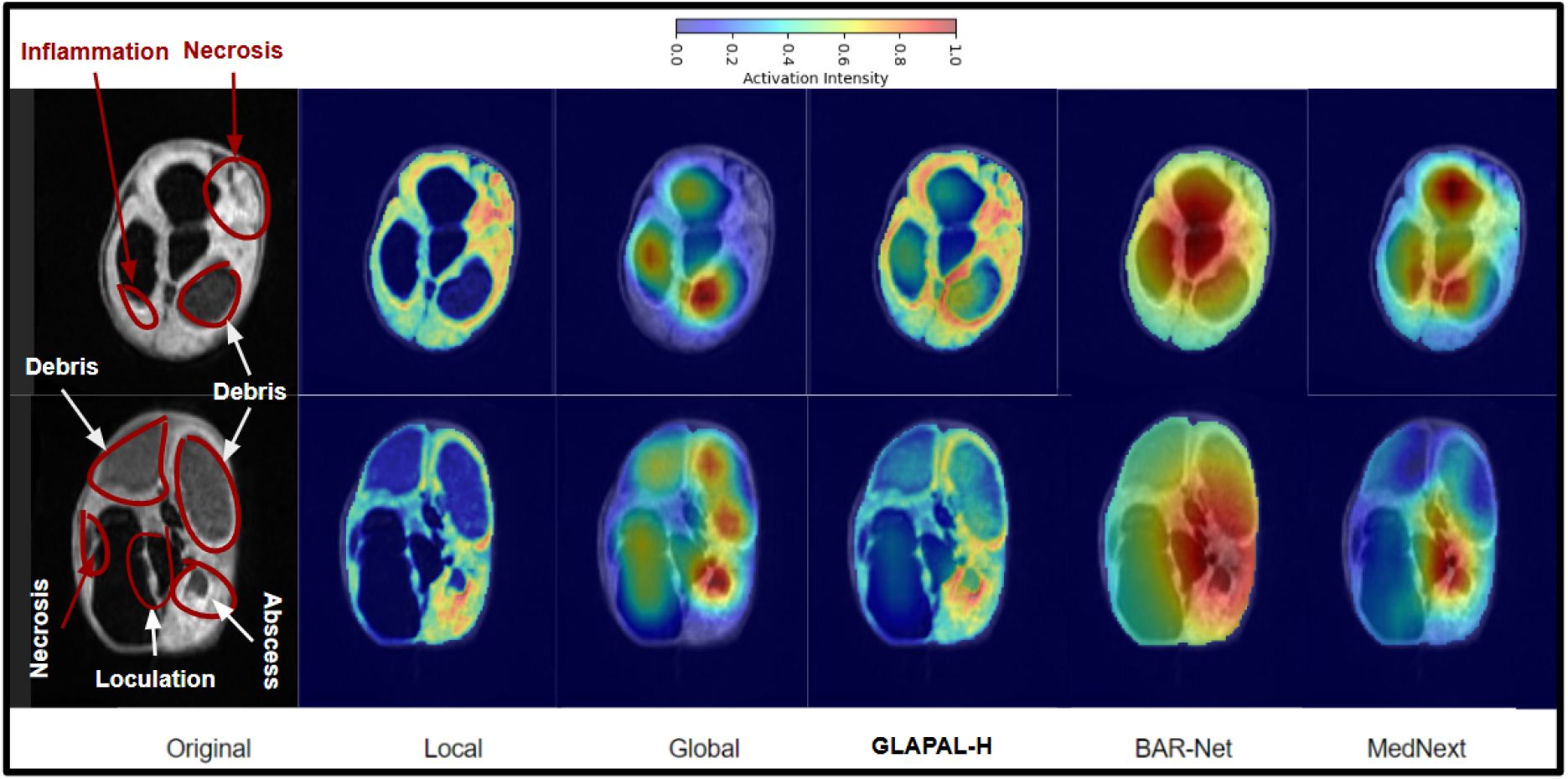
Class Activation Maps (CAM) of two PIH infants corresponding to the local and global branches of GLAPAL-H, CAM of GLAPAL-H, along with CAM of MedNeXt and BAR-Net for the Three Class Problem.

### F Edge Case Performance

Edge cases, such as the misclassified examples illustrated in Fig. 8, highlight the inherent diagnostic challenges in distinguishing hydrocephalus etiologies, particularly when using low-field MRI. Visual clinical assessment relies on visual manifestations of infection, such as large-scale global asymmetry and local signs like septation, abscesses, or debris, as shown in Figs. 1 , 7. However, in challenging edge cases like those shown in Fig. 8, these visual manifestations of infection can be subtle, overlap between classes, or be misinterpreted, leading to diagnostic uncertainty and false positives or false negatives in all the models. For instance, the PIH case misclassified as NPIH presents with relatively symmetric ventricles and lacks strong visual cues for local infections, potentially resembling NPIH. Conversely, the NPIH case misclassified as PIH shows localized bright signs of infection, which the model might incorrectly interpret as a sign of local infection due to high contrast regions, despite the absence of PIH etiology. While GLAPAL-H demonstrates superior overall performance and robustness compared to state-of-the-art methods, these edge cases underscore the limitations of current methods on low-field MRI. We conjecture that the integration of additional pulse sequences, such as T2-weighted and proton density imaging, could further mitigate diagnostic ambiguities by providing complementary perspectives. Such multi-modal approaches may offer a richer depiction of infectious and inflammatory processes, potentially improving the differentiation of subtle visual cues, as elaborated next.

**Fig. 8.**
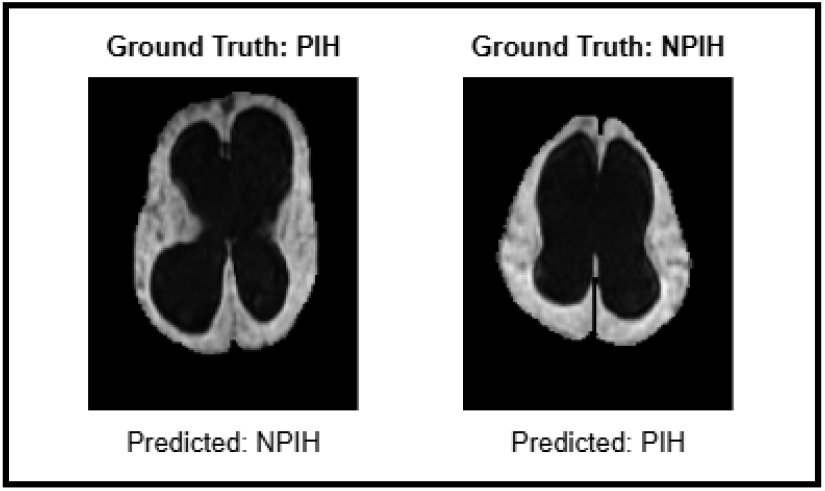
Examples of Misclassified Edge Cases.

## VI. Discussions and Future Work

### Limitations

This study is limited by the relatively small dataset from a single institution. Expanding the number of contributing institutions will be critical for increasing the dataset size for both postinfectious and non-postinfectious hydrocephalus cases. Regional differences in infectious agents will introduce variability, as patterns of postinfectious hydrocephalus differ by geography. Fundamentally, postinfectious hydrocephalus represents an inflammatory response to infection and falls within the broader classification of post-inflammatory hydrocephalus [77]. GLAPAL-H will require further training on this expanded and more diverse dataset to generalize across the broader spectrum of infectious patterns.

### Clinical Validation

Infectious validation represents a critical challenge in imaging-based diagnostics. In regions where specific organisms dominate infectious etiologies and postinfectious imaging patterns, targeted diagnostic approaches, such as organism-specific PCR, enable effective validation of ongoing or recent infection. However, in most settings, broader validation encompassing all nonhuman genomes within a sample is required. This necessitates an unbiased pathogen discovery approach, typically through metagenomic strategies, which remain an active focus of current research. For near-term purposes, GLAPAL-H is being validated against expert clinical and imaging assessments of infection, serving as a proxy until broader metagenomic validation becomes feasible.

### Future Work

In future work, infections should be characterized not by static snapshots but by the evolving trajectory of pathogen–host interactions. In brain infections, two critical clinical questions arise: (1) whether the infection remains active and necessitates ongoing antimicrobial therapy, and (2) during treatment, whether imaging can guide clinicians in determining the optimal timing for discontinuing antimicrobial agents. A related challenge is whether maladaptive inflammatory responses can be pharmacologically modulated to prevent or mitigate hydrocephalus while antimicrobial treatment is administered. Point-of-care imaging modalities, such as those studied here, offer immediately accessible information. With the increasing global dissemination of low-field MRI, such imaging represents a sustainable resource for clinical decision-making across both high- and low-resource settings. Future approaches that integrate multiparametric pulse sequence data (e.g., T1, T2, FLAIR, proton density, diffusion) will enable independent assessments of the chemical microenvironment of brain structures. The fusion of these complementary contrasts may yield a richer, more nuanced depiction of infectious and inflammatory processes, supporting more precise and individualized management. Future work could also integrate advanced interpretability methods, combining vision and language in a multi-modal framework with image data, along with radiology reports to align the visual and textual representations of infection signs. This will enhance a trustworthy human-AI collaboration for hydrocephalic patient care.

## V. Conclusion

Our key contributions include the development of GLAPAL-H, a new domain-enriched architecture that leverages local and global branches with parts-based guidance to capture infection signs in hydrocephalic patients. By introducing Two Class and Three Class problems alongside a low-training setup, we demonstrate GLAPAL-H’s robustness and reliability under challenging conditions, while showing the enhanced interpretability of GLAPAL-H using the trinary activation maps compared to the SOTA methods. In the Two Class Problem, GLAPAL-H outperforms BAR-Net despite using low-field MRI instead of high-quality CT scans, highlighting its potential for cost-effective, radiation-free infection diagnosis in resource-constrained settings.

## Data Availability

All data produced in the present study may be available upon reasonable request to the authors, although there are restrictions on personal health information on the subjects.

## Footnotes

1 Henceforth, for ease of exposition, we refer to low-field FLAIR MRI as just FLAIR.

