## Supplementary Figure for "GLAPAL-H: Global, Local, And Parts Aware Learner for Hydrocephalus Infection Diagnosis in Low-Field MRI"

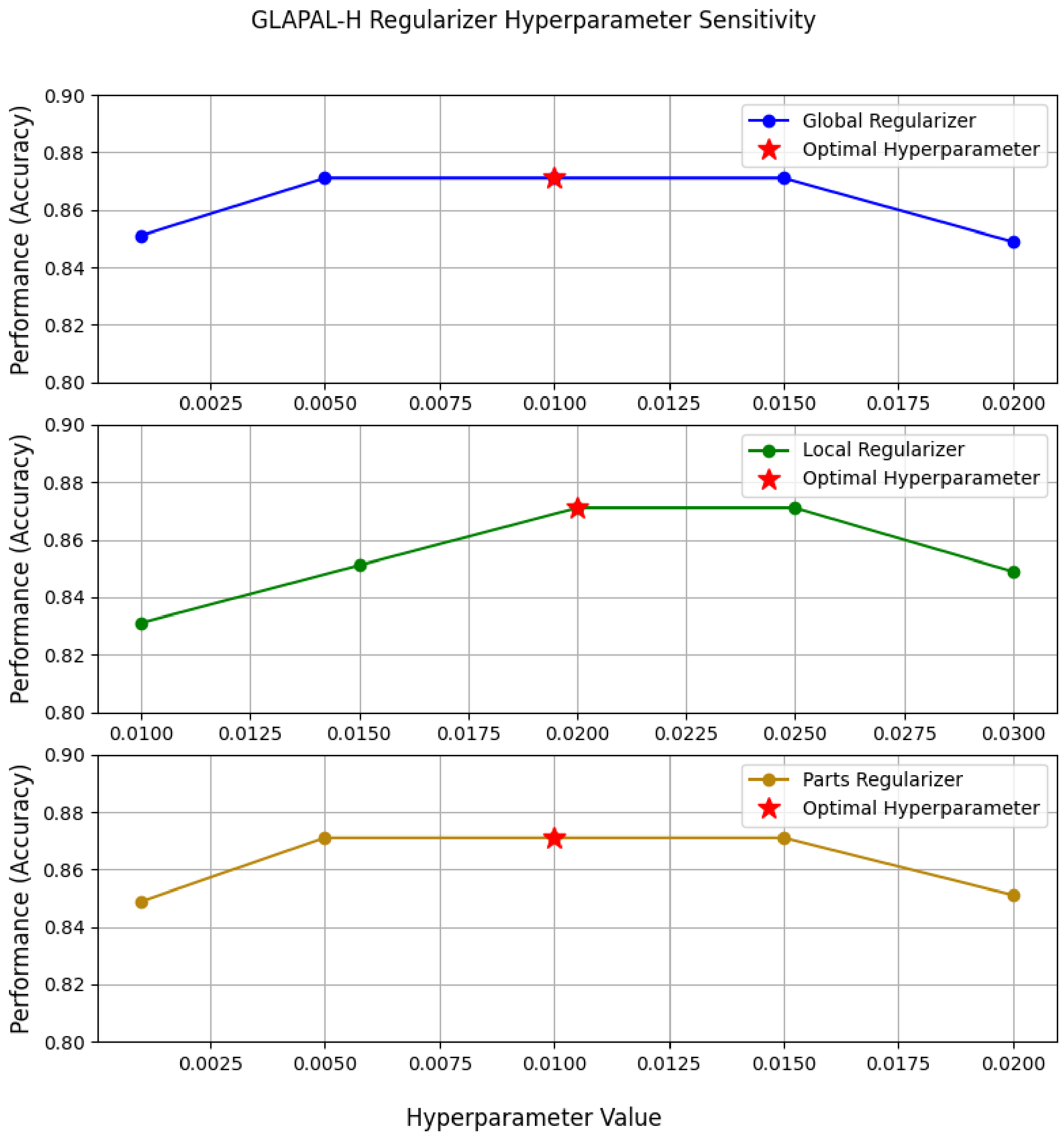


Fig. S-1. Hyperparameter sensitivity analysis of the Global, Local, and Parts regularizers in the vicinity of the aforementioned optimal hyperparameter values (0.01, 0.02, 0.01) for Global, Local, and Parts regularizers, respectively. When varying one hyperparameter, others were kept fixed to their optimal values. The results above unequivocally confirm that under small perturbations around optimal hyperparameter choices GLAPAL-H exhibits desirable robustness for practical deployment.
